# Sex-dependent manifestations of intracranial aneurysms

**DOI:** 10.1101/2023.03.29.23287441

**Authors:** Thomas Wälchli, Martin Ndengera, Paul E. Constanthin, Jeroen Bisschop, Sandrine Morel, Oliver Gautschi, Moncef Berhouma, Aristotelis Kalyvas, Philippe P. Monnier, Ethan A. Winkler, Hans Kortman, Kartik Bhatia, Philipp Dammann, Max Jägersberg, Renato Gondar, Karl Schaller, Brenda R. Kwak, Philippe Bijlenga

## Abstract

**Background:** Intracranial aneurysms (IAs) are more common in females than in males, however, there is still very limited knowledge on sex-dependent differences regarding aneurysm location, multiplicity, rupture risk, risk factors and histopathology.

**Methods:** This prospective, consecutive cohort study examined whether IAs differ in multiplicity, location, geometry, rupture risk, risk factors and histology between sexes.

**Results:** We included 982 patients (714 women, 268 men) totaling 1484 IAs (1056 unruptured, 397 ruptured). Three hundred sixty-three patients (36.97%) had multiple IAs, the proportion of which was significantly higher in females. In women, the ICA (40.79%) was the most frequent location for IAs, whereas in men most were found along the ACA territory (32.86%). Men were significantly more often diagnosed with ruptured aneurysms. Aneurysm geometry did not differ between sexes, however, ruptured aneurysms in men presented with a significantly larger neck diameter than unruptured ones. Regarding risk factors for aneurysm rupture, blood pressure control was more effective in women, whereas the effect of smoking status did not show clear sex-dependent differences. Histologically, wall-type classification analysis showed significantly more severe aneurysm wall types in men.

**Conclusion:** IA prevalence in women is significantly higher than in men. Women more often present with multiple IAs whereas men were more often diagnosed with ruptured IAs. Sex-specific differences in IA location were identified whereas geometry of IAs did not differ between sexes. IAs in men showed a more severe histological wall type. Further research is needed to unravel the molecular mechanisms underlying these important sex-dependent manifestations in IAs.

## Introduction

Intracranial aneurysms (IAs) are focal weakenings of cerebral arteries present in the subarachnoid space^1,2^ with a general prevalence of 2-4%^3,4^. IA rupture results in aneurysmal subarachnoid hemorrhage (aSAH) with an annual incidence of 6-8/100,000/year in Western countries^2,5–7^, leading to significant mortality and morbidity.^2^ Known risk factors for the development of IAs are smoking, hypertension, alcohol, hypercholesterolemia, estrogen deficiency, carotid artery stenosis and familial history of aneurysms.^8^ Genetic (familial) risk factors are both inherited and acquired. Inherited genetic risk factors encompass single gene mutations (8q^9,10^, 9p^9,10^, 4q^10^) or genetic diseases such as polycystic kidney disease, Ehlers-Danlos-, Loeys-Dietz- and Marfan syndrome, hereditary hemorrhagic telangiectasia and neurofibromatosis type 1^11,12^. Acquired genetic risk factors refer to genetic mutations resulting in a higher chance of IA development and/or rupture upon smoking and alcohol consumption (e.g. single nucleotide polymorphisms on chromosome 9p21).

According to the International Study of Unruptured Intracranial Aneurysms (ISUIA), symptoms related to unruptured IAs (UIAs) are rare for IAs until 7 mm^13,14^ and uncommon for a maximum diameter of 8-20 mm^13^. When present, symptoms related to cranial nerve compression (ocular motor palsy, visual field defects), epilepsy and cerebral infarction are the most frequent^15,16^. With the multiplication of imaging facilities and increased technical sensitivity, the rate of diagnosis of incidental UIAs is increasing and currently around 3% in the middle-aged population^4,17^.

The decision to treat UIAs depends on the “risks-benefits”-balance of treatment options (neurosurgical or endovascular) versus the annual (cumulative) rupture risk and is evaluated on a case-to-case basis^2,13^. Preventive treatment exposes patients to a 1% mortality and 5% morbidity risk^18–20^. Known risk factors for IAs rupture partially overlap with developmental risk factors and encompass other parameters such as size and location^8^. The Japanese Unruptured Cerebral Aneurysm Study (UCAS) showed that aneurysm size influences the rupture risk. Moreover, they showed that posterior circulation IAs ruptured more often than aneurysms in the anterior circulation (anterior (ACA) and middle cerebral arteries (MCA))^21^. Of note, the published PHASES score for prediction of rupture risk included the parameters Population, Hypertension, Age, Size, Earlier aSAH and Site (location)^14,22,23^.

Understanding the natural history of IAs is imperative to optimally treat UIAs and sex-differences have only been addressed by a limited number of dedicated studies, consisting predominantly of ruptured aneurysms. In a cohort of 906 patients with ruptured IAs, Kongable and colleagues reported that women have a 3-times higher incidence of IAs (more often multiple) and a higher proportion of cavernous and internal carotid artery (ICA) IAs, while men more often had ACA IAs^24^. Similar findings were reported in a cohort totaling 682 ruptured and unruptured aneurysms, with a higher incidence of aSAH among women^25^. However, due to a lower prevalence difference, the latter might disappear when studying case-matched controls and future research is needed. Of note, the existence of sex differences regarding IA rupture still remains debated due to contradictory results between more recent studies^26–28^ and little is known regarding the influence of sex on IA biology, particularly regarding risk factors for rupture and histopathology.

Therefore, the aim of this study was to investigate sex-dependency of IAs with regard to (1) multiplicity and location (2) rupture risk and size/morphology, (3) established risk factors and (4) histological properties of the aneurysm wall.

## Materials and methods

### Study design

This study analyses prospectively collected data from patients recruited at the Geneva University Hospital (HUG). Registry enrolment began on November 1^st^ 2006. Data were extracted on October 31^st^ 2017. Registry details and standardization of data collection were described previously^14,29^.

### Patient population

Patients aged ≥18y.o. and diagnosed with both ruptured and unruptured IAs based on MR angiogram (MRA), high-resolution three-dimensional CT angiogram (3D-CTA), digital subtraction angiography (DSA) or digital rotation angiography (DRA) imaging were eligible for inclusion. For detailed imaging characteristics, we refer to Gondar et al.^29^. Exclusion criteria were: 1) suspicion of IA(s) on MRA or CT-angiography that were not confirmed angiographically, 2) extradural, intracranial aneurysms, 4) missing or inconsistent medical data, 5) refusal for e-health records to be used for research purposes and 6) incomplete consent.

### Data collection

Basic characteristics (Supplementary Tables 1 and 2) including patient history, risk factors for IA development and rupture (including smoking (≥100 cigarettes during life-time) and hypertension (>140/90 mm Hg)), as well as neuroradiological details (IAs number, location (according to the @neurIST nomenclature^30^) and morphology (saccular on bifurcations, saccular on sidewall or fusiform)) were collected for all recruited patients. MRA, (3D)-CTA, DSA and/or DRA were clinically routinely reviewed by two independent clinicians (one vascular neuroradiologist and one vascular neurosurgeon). Aneurysm neck and maximum dome diameters and morphological information (irregularities, blebs or lobules) were documented (Supplementary Table 1).

### Ethical approval

All procedures performed in studies involving human participants were in accordance with the ethical standards of the institutional and/or national research committee and with the Helsinki declaration. Written informed consent was obtained from all participants and the HUG had institutional review board approval (@neurIST protocol, ethics authorization PB_2018-00073, previously CCER 07-056). All data were deidentified.

### Histological analysis

The AneuX - Intracranial Aneurysm Domes biobank has been certified by the Swiss Biobanking Platform (SBP) with a VITA quality label (certificate number HUG_2202_6). Saccular IA samples were collected by resecting the aneurysmal dome after neurosurgical clipping. Domes were fixed in formaldehyde and stored at room temperature. After being embedded in paraffin, aneurysm domes were sectioned at 5 μm thickness and stained with hematoxylin-eosin or immunolabelled with antibodies allowing the visualization of vascular smooth muscle cells (vSMCs) and endothelial cells as previously described^31^. Aneurysm walls were then classified accordingly to Frösen et al.^32^: A: endothelialized wall with linearly organized vSMCs; B: thickened wall with disorganized vSMCs; C: hypocellular wall; D: extremely thin hypocellular wall. Each dome received two scores, a first characterizing the dominant wall type observed in the resected sample (covering the majority of the dome surface) and a second being representative of the most severe focal histological^31^.

### Statistical analysis

GraphPad Prism version 9.3.0 for macOS (GraphPad Software, San Diego, California USA) was used for statistical analysis and graphical data representations.

The parameters obtained from female and male populations were compared using Mann-Whitney *U*-tests. Comparisons of differences between groups were performed with 2-tailed, unpaired T-tests (continuous variables) and Chi-Square-tests (categorial variables). 2-way ANOVA with Bonferroni posthoc-analysis was used to compare several groups. A p-value <0.05 was considered statistically significant. Data are presented as mean ± (SD), median ± (SD) or as proportions (%). Detailed statistical values (exact p-values, Odds Ratio’s (ORs) and SD) are indicated in the figure legends. For all quantitative analyses, subdivided data sets with less than 10 patients (cumulative men and women) were pooled together and referred as “other”.

### Data availability

Raw data were generated at the Geneva University Hospital. Derived data supporting the findings of this study are available from the corresponding author on request.

## Results

### Patients’ characteristics

A total of 1442 patients were diagnosed with IAs either incidentally (no or non-related symptoms) or upon aSAH. Of those, 460 (31.9%) were excluded according to exclusion criteria leaving 982 patients (714 women and 268 men) for analysis, representing 1484 IAs (1113 in females and 371 in males) (Figure 1).

**Figure 1.**
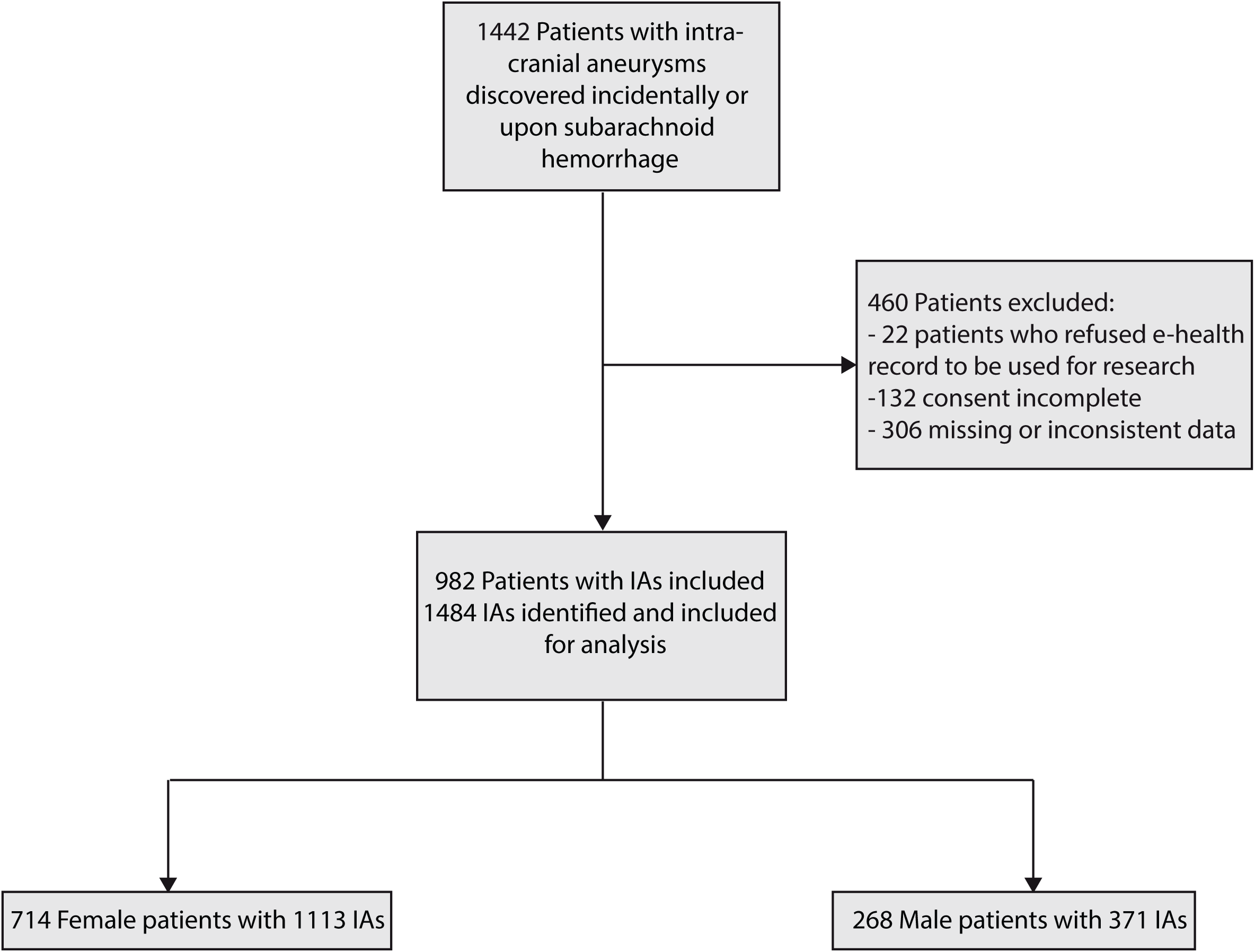
Flow diagram indicating the number of patients who were screened and included in the analysis. The different sections of the protocol are indicated in grey boxes. Patients with diagnosed IAs were screened for eligibility in the analysis. After applying pre-defined exclusion criteria, 714 female patients and 268 male patients were included in the analysis.

### Women present with both more single and multiple IAs than men

First, we investigated the distribution of IAs between sexes. Among the 982 patients (mean age [±SD], 55.8 [±13.96] years), 714 (72.71%) were women (56.1 [±13.39] years) and 268 (27.29%) were men (55.6 [±14.06] years) (P<0.0001). They presented a total of 1484 IAs, which were significantly more numerous (1113 versus (vs) 371) and showed a higher proportion (75% vs 25%) in females (P<0.0001; OR=9.08) (Figure 2a-c).

**Figure 2.**
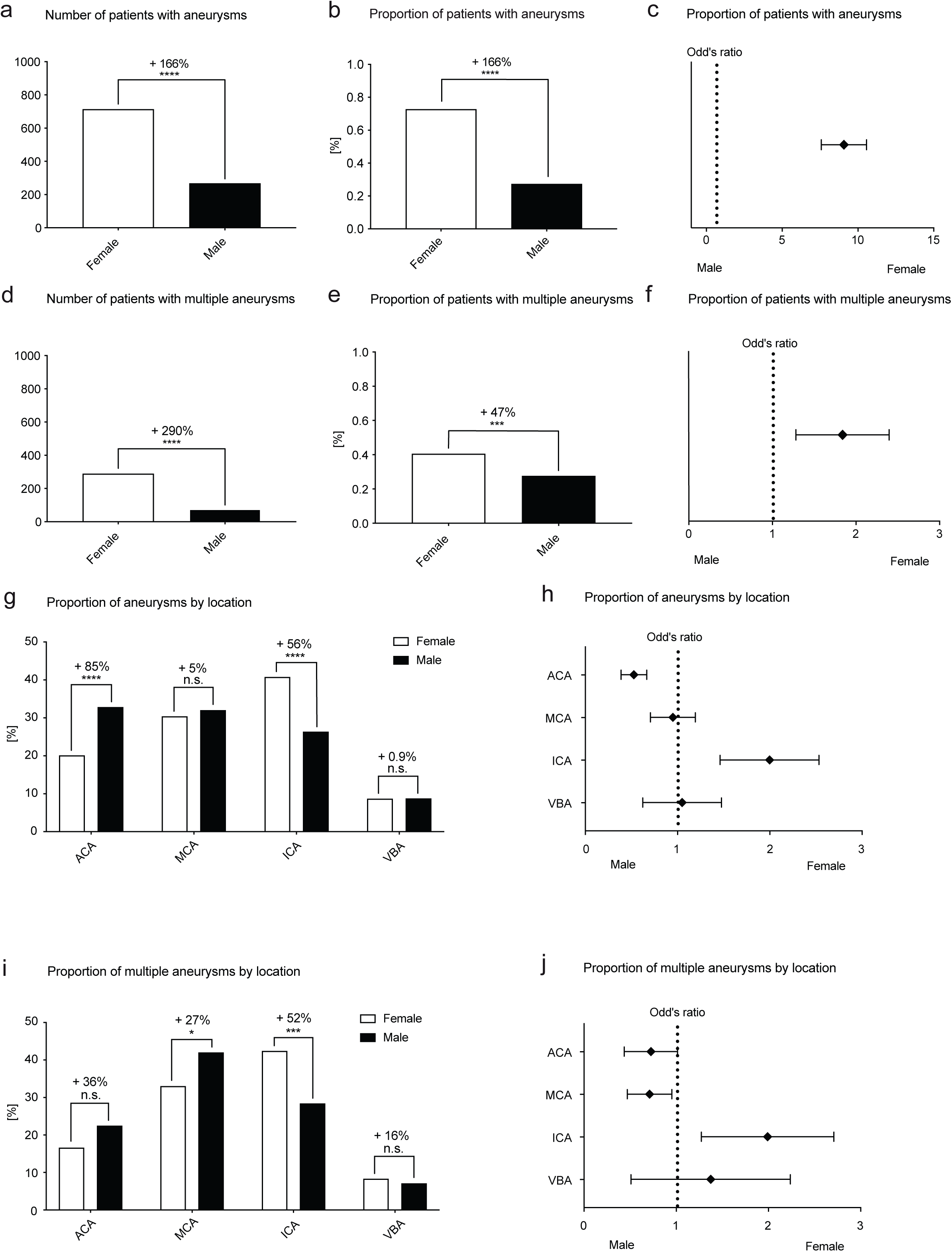
Women have more and more often multiple intracranial aneurysms than men. **A-F** Significantly more patients were female than male (women n = 714 (72.71%), men n = 268 (27.29%); P < 0.0001; OR 9.08) (**A-C**). Women have significantly more aneurysms (women n = 1113 (75.0%), men n = 371 (25.0%)) and more often multiple aneurysms than men (multiple IAs: women n= 289/714 (40.48%), men = 74/268 (27.61%); P = 0.0002; OR = 1.783) (**D-F**). **G,H** Proportion of aneurysms by location. The proportion of ACOM aneurysms was significantly higher in male as compared to female (+83%; P < 0.0001; OR = 2.217) whereas ophthalmic segment carotid artery aneurysms and PCOM aneurysms were significantly more frequent in female as compared to male (+146%; P < 0.0001; OR = 2.782) (**G,H**). **I,J** Proportion of multiple aneurysms by location. The proportion of ophthalmic segment carotid artery aneurysms was significantly more frequent in female as compared to male among the patients with multiple IAs (+244%; P < 0.0001; OR = 3.91) (**I,J**). *P < 0.05, **P < 0.01, ***P < 0.001, ****P < 0.0001 (Fisher exact test).

Next, we addressed the proportion of patients with multiple aneurysms. Interestingly, 289 (40.48%) female and 74 (27.61%) male patients presented with multiple aneurysms resulting in a significantly higher proportion in women (P=0.0002; OR=1.783) (Figure 2d-f). Together, these results show that the overall prevalence of aneurysms, both single and multiple, is higher in women.

### IA localization is sex-dependent

Next, we compared the differences in localization of IAs between female and male patients by applying the Koivisto chart system (commonly utilized since 2000)^33^ to our cohort. We also provide the exact location of single and multiple IAs for both sexes.

In women, the MCA bifurcation (20.95%) was the most frequent location, followed by the ICA ophthalmic segment (OICA) (16.74%), the anterior communicating artery (ACOM) (14.97%), the posterior communicating artery (PCOM) (10.10%) and the MCA M1 segment (7.39%). Aneurysms in men showed a different distribution with ACOM (27.48%) being the most frequent location, followed by the MCA bifurcation (20.68%), M1 (8.22%), OICA (6.80%) and PCOM (6.80%). When compared by localization, the proportion of ACOM aneurysms was significantly higher in men (27.48%) than in women (14.97%) (P<0.0001; OR=2.217) whereas the proportion of OICA aneurysms was significantly higher in women (16.74%) than in men (6.79%), (P<0.0001; OR=2.782). PCOM aneurysms showed a non-significant trend to be more abundant in women (10.10%) than in men (6.80%) (P=0.0581; OR=1.555). No other significant difference was observed (Figure 2g-h).

Regarding multiple IAs, OICA aneurysms were significantly increased in women (16.29%) compared to men (4.73%; P<0.0001; OR=3.91) (Figure 2i,j). No other significant difference was found (Figure 2i,j, Supplementary Figure 1).

### Taken together, these results reveal sex-specific differences in IA location with a predominance of ACOM aneurysms in men and of OICA aneurysms in women

### IAs tend to rupture more frequently in men

IA rupture results in aSAH with possible concomitant intracerebral hemorrhage, leading to significant mortality and morbidity^34^. We therefore investigated the number and proportion of ruptured aneurysms in both sexes. Whereas the absolute number of ruptured aneurysms was higher in women, the proportion of ruptured aneurysms (32.78%) was significantly higher in men (25.50%; P<0.01; OR=1.425) (Figure 3a-c). Moreover, aneurysms of the ACA showed significantly more ruptures in men (45.61%) than in women (34.94%; P<0.05; OR=1.561) (Figure 3d,e). No significant difference was observed for other locations (Figures 3d,e, Supplementary Figures 1 and 2).

**Figure 3.**
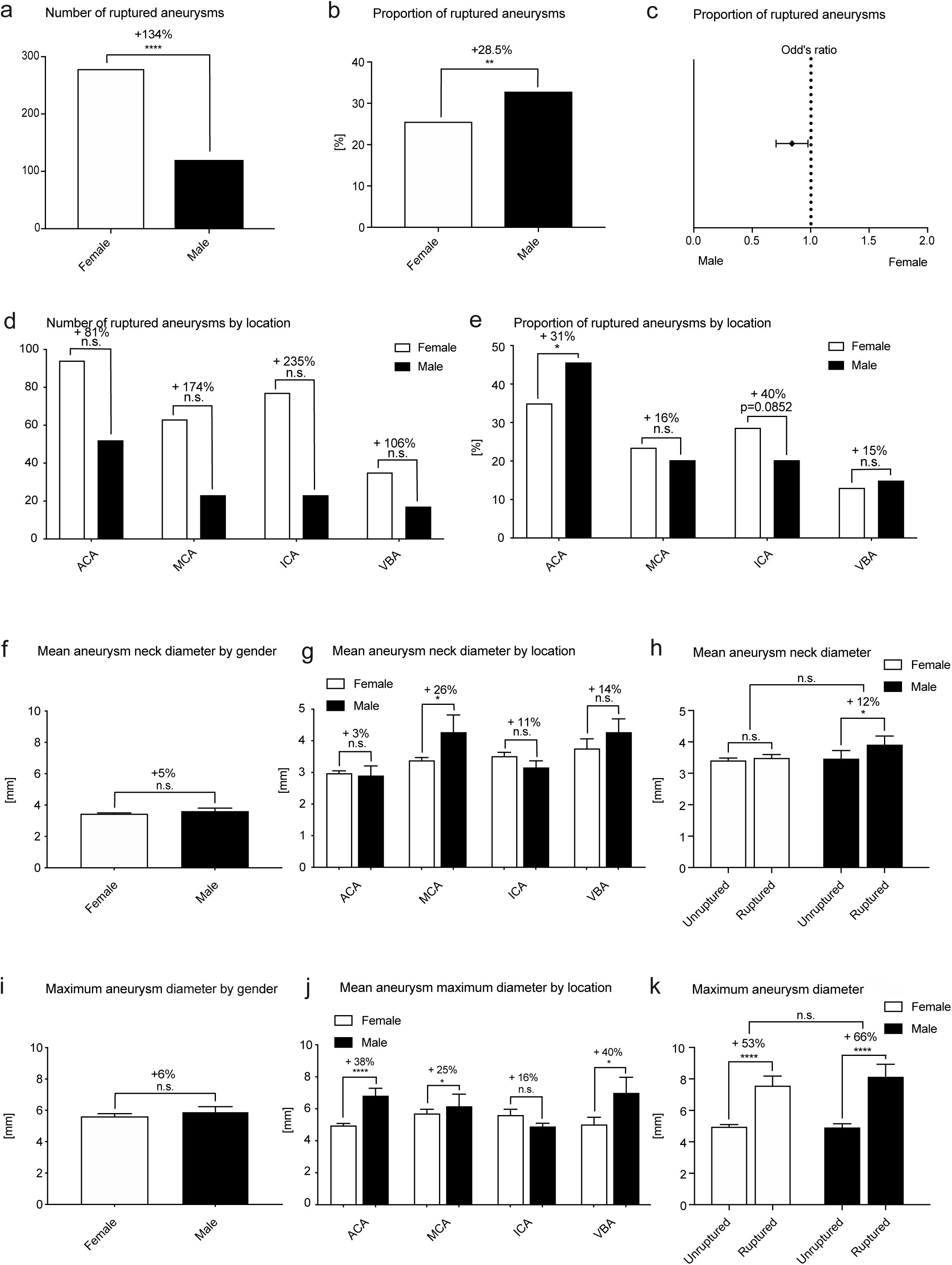
Intracranial aneurysms rupture more frequently in men than in women, depending on neck (but not maximum) aneurysm diameter. **A-C** Whereas the absolute number of ruptured aneurysms was higher in women (n=278) than in men (n=119) (**A**), the proportion of men diagnosed with a ruptured aneurysm (119/363 = 32.78%) was significantly higher than women (278/1090 = 25.50%; P<0.01; OR = 1.425) (**B,C**). **D,E** Analysis of the number and proportion of ruptured aneurysms by location revealed a significantly higher proportion of ruptured aneurysms in the ACA territory in males (52/114 = 45.61%) than in females (94/269 = 34.94%; P<0.05; OR = 1.561). **F-K** Intracranial aneurysm geometry as revealed by neck diameter (**F-H**) showed a significantly lower neck diameter in the MCA territory for females (mean 3.378 mm) than in males (4.269 mm; P<0.05) and significantly higher maximum IA diameter in the ACA and MCA territories for males versus females (ACA P<0.0001; MCA P<0.05, respectively) (**I-K**). Neck diameter was significantly higher in ruptured aneurysms as compared to unruptured aneurysms in men but not in women (ruptured: 3.91 [3.89] mm; unruptured: 3.467 [2.85] mm; +12%; P<0.05) (**H**). Maximum IA diameter was significantly larger in ruptured IAs as compared to unruptured IAs, for both women (ruptured: 7.571 [9.491] mm; unruptured: 4.956 [3.887] mm; P<0.0001) and men (ruptured: 8.131 [8.108] mm; unruptured: 4.905 [3.689] mm; P<0.0001) (**K**). *P<0.05, **P<0.01, ***P<0.001, ****P<0.0001 (Fisher exact test).

### Taken together, these results suggest that IAs rupture more often in men, particularly ACA aneurysms

### IAs size differences between men and women

Next, we addressed the size of IAs in sexes by comparing maximum IA diameter and mean neck diameter. Mean neck diameter did not differ between women (3.442 [2.113] mm) and men (3.607 [3.579] mm) (P=0.3056) except for MCA IAs that showed larger mean neck diameters in males (Figure 3f,g). Similarly, the overall maximum aneurysm diameter did not differ between women (5.604 [5.896] mm) and men (5.924 [5.664] mm) (P=0.3886), but was significantly higher in the ACA, MCA and VBA in males (Figure 3i,j). When comparing ruptured and unruptured aneurysms, there were no significant differences regarding aneurysm mean neck diameter in women. Ruptured aneurysms in men, however, presented with a significantly higher mean neck diameter (ruptured: 3.91 [2.85] mm; unruptured: 3.467 [3.891] mm; P<0.05) (Figure 3h). The maximum aneurysm diameter was significantly higher in ruptured aneurysms for both women (ruptured: 7.571 [9.491] mm; unruptured: 4.956 [3.887] mm; P<0.0001) and men (ruptured: 8.131 [8.108] mm; unruptured: 4.905 [3.689] mm; P<0.0001) (Figure 3k, Supplementary Figures 1 and 2).

These data reveal that the maximum aneurysm diameter is significantly higher in ruptured aneurysms as compared to unruptured aneurysms in both women and men. However, mean neck diameter of ruptured aneurysms was significantly higher only in men suggesting that aneurysmal size and morphology might be more predictive for the rupture risk in men.

### Quitting smoking and treating high-blood pressure is more effective in women

Smoking and hypertension are well-known risk factors for IAs^34^. Therefore, we next sought to investigate these risk factors in our patient cohort.

To investigate the effects and possible sex-dependent differences of smoking, patients were categorized into: 1) patients who never smoked (“non smoking”), 2) patients that stopped smoking before the diagnosis of IA (“quit smoking”) and 3) patients still smoking at the time of the diagnosis (“still smoking”). In out cohort, men smoked more frequently (Figure 4a). Furthermore, when comparing ruptured with unruptured aneurysms for both sexes according to smoking status, the proportion of patients with ruptured aneurysms who quit smoking was significantly higher for men (Figure 4c-e and Supplementary Figure 3a-f).

**Figure 4.**
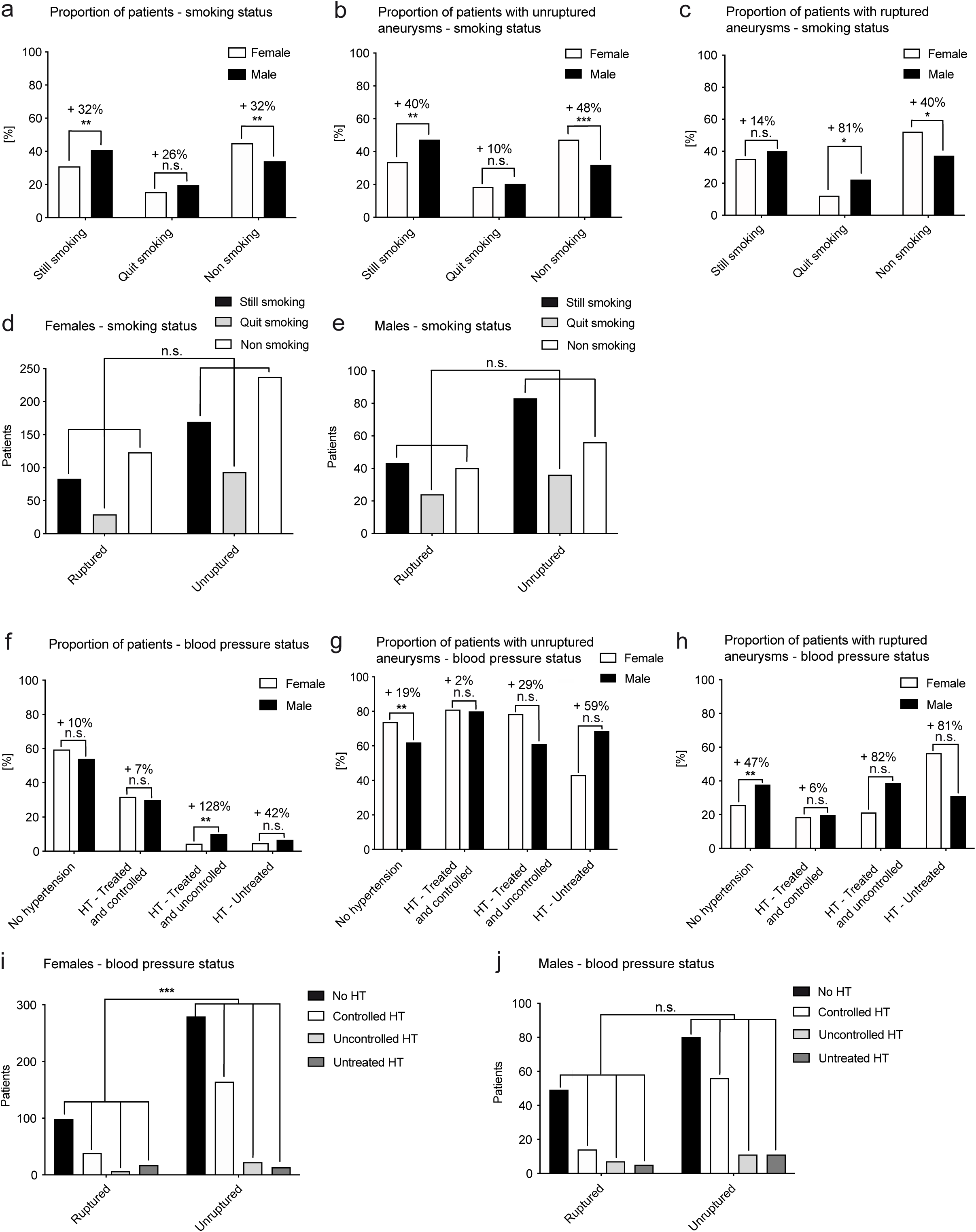
Male patients affected by IA smoke more frequently than female. **A-D** Patients with IAs were classified, according to smoking status, into “still smoking”, “quit smoking”, and “non-smoking” Overall, there were no differences in smoking status among male and female patients (**A**). The proportion of smokers as compared to non-smokers and divided into patients with ruptured and unruptured IAs did not show differences in females (**B**) nor in males (**C**). **D-F** Patients with IAs were classified, according to blood pressure, into “no hypertension”, “treated and controlled hypertension”, “treated and uncontrolled hypertension” and “untreated hypertension”. Overall, there were no significant differences in blood pressure status among male and female patients (**D**). When specified for patients with unruptured aneurysms (**E**) and ruptured aneurysms (**F**), no significant sex-dependent changes were found. **G-I** When comparing more specifically, significantly fewer ruptured aneurysms were observed among women with treated hypertension (P<0.001) (**H**) and normotension (P<0.01) (**I**) as compared to women with untreated hypertension. **J-L** In men, patients with treated hypertension presented with a significantly higher proportion of ruptured aneurysm as compared to patients without hypertension (P<0.05). *P<0.05, **P<0.01, ***P<0.001 (Fisher exact test).

Next, regarding blood pressure (BP) and its effects and possible sex-dependent differences in our cohort, patients were categorized into: 1) normotension, 2) treated-and-controlled hypertension, 3) treated-and-uncontrolled hypertension, and 4) untreated hypertension. In our cohort, no differences were found between men and women with regard to the overall BP status (Figure 4f). Among the patients with unruptured IAs, there were significantly more females without hypertension whereas among patients with ruptures IAs, the proportion of patients without hypertension was significantly higher in males (Figure 4g,h). Interestingly, for ruptured aneurysms in women, the proportion of women with treated hypertension was similar to the proportion of women without hypertension. Accordingly, significantly fewer ruptured aneurysms were observed among women with treated hypertension and normotension as compared to women with untreated hypertension (P<0.0001 and P<0.001, respectively), indicative of the benefit of this treatment modality. In men, on the other hand, patients with treated hypertension still presented with a significantly higher proportion of ruptured aneurysm as compared to patients without hypertension (P<0.05) (Figure 4i-j and Supplementary Figure 3g-l), suggesting a less important effect of treatment of hypertension. Taken together, these results indicate that sex-differences exist regarding IA rupture risk factors, with women benefitting significantly more from treatment of hypertension while men do not appear to benefit from it.

### Aneurysm walls show a more severe pathological phenotype in men

We next addressed histopathological features among sexes. We analyzed the histology of 48 (12 males and 36 females) saccular aneurysm walls and classified them according to the classification proposed by Frösen et al^32^. In this classification system, aneurysmal walls are classified into four categories, depending on their cellular components, with A being the least “severe” wall type and D being the most “severe” wall type^32^ (Figure 5a-l). Each aneurysm was graded twice (“dominant” and “most severe” wall type grades). Referring to the dominant wall type scoring, 11%, 44%, 37% and 8% of the resected domes were classified as wall type A, B, C and D in women whereas 8%, 34% and 58% were classified as wall type A, B and C in men, demonstrating a significant difference in the distribution of histological severity grades between women and men (Figure 5m-n). In women, the dominant wall type being most present was the wall type B, whereas in men, the majority of the aneurysm domes were classified as wall type C (Figure 5m-n). Referring the most severe focal histological wall type grading (Figure 5o-p), we found that 100% of the resected domes were classified as wall type C or D, *i.e.* degenerative histological phenotypes, in men, in comparison to 72% in women (Figure o-p).

**Figure 5.**
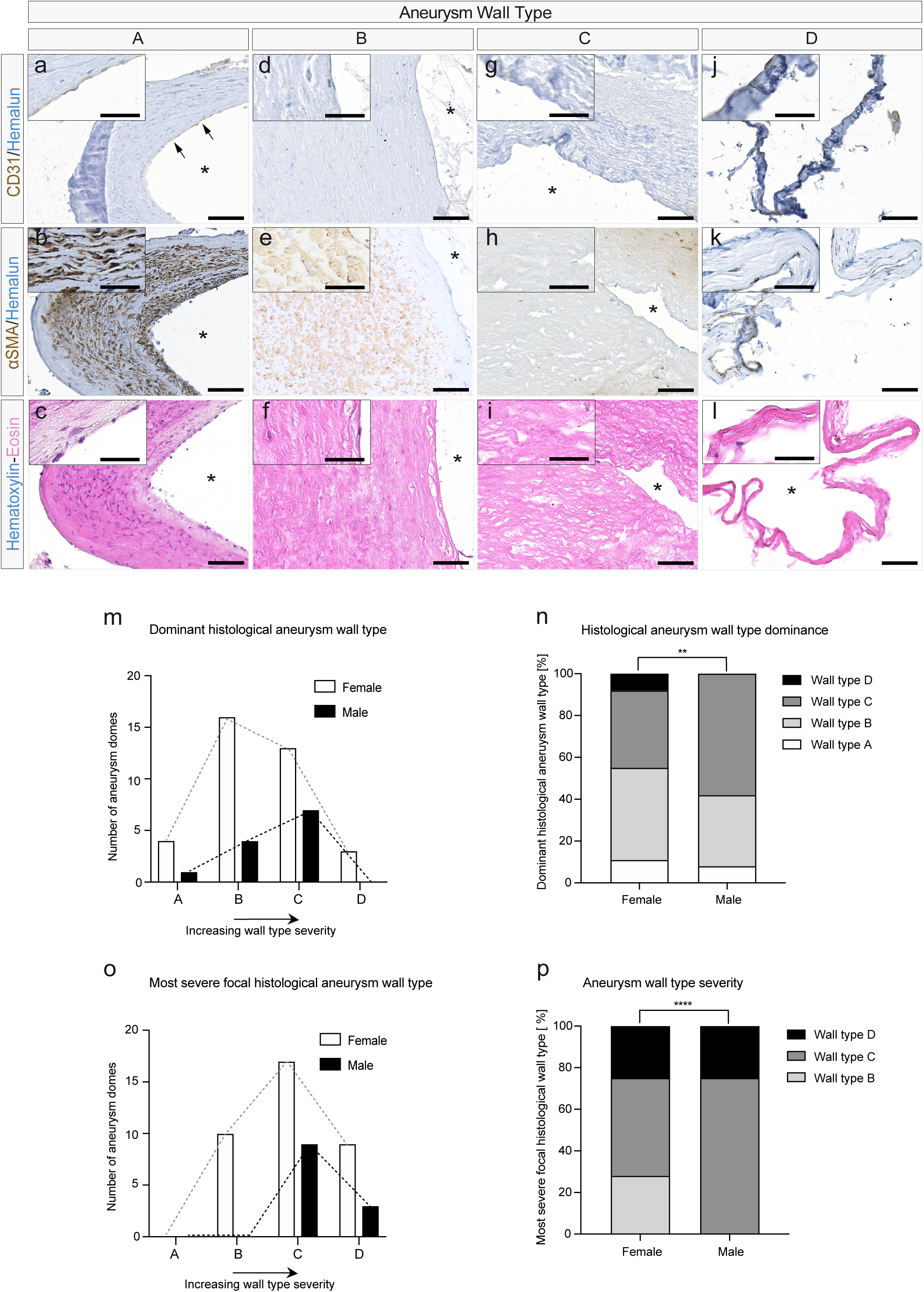
Histological classification of aneurysm wall type severity. **A-L** Representative examples of stained intracranial aneurysm dome sections allowing aneurysm wall histological classification. **A-C** Wall type A is characterized by an endothelialized wall (CD31: endothelial cell staining, some endothelial cells are indicated by arrows) with linearly organized smooth muscle cells (α-sma: α-smooth muscle actin staining, in brown). **D-F** Wall type B shows a thickened wall with disorganized vascular smooth muscle cells. **G-I** Wall type C shows characteristic hypocellularity. **J-L** Wall type D is extremely thin and hypocellular. Total cellular content is shown with the Hematoxylin-eosin (HE) staining (**C, F, I, L**). **M-P** Quantification was performed by characterization of the dominant histological wall type within the individual samples (**M, N**) (11%, wall type A; 44%, wall type B; 37% wall type C; and 8%, wall type D in females, and 8%, wall type A; 34%, wall type B; 58% wall type C; and 0%, wall type D in males), and based on the most severe focal histological wall type (**O, P**) (0%, wall type A; 28%, wall type B; 47% wall type C; and 25%, wall type D in females, and 0%, wall type A; 0%, wall type B; 75% wall type C; and 25%, wall type D in males). Chi-square test, **P<0.01; ****P<0.001; scale bars, 100 μm (**A-L**, overviews); 50 μm (**A,L**, insets). Asterisks represent the luminal side of the aneurysm.

Taken together, these results indicate that aneurysms in men exhibit histopathological features that correlate with a higher risk for IA rupture..

## Discussion

Here, we report on sex-dependent differences observed in a prospective, consecutive cohort study including 982 patients with 1,484 IAs. We show that women are more often diagnosed with IAs and that IA location and multiplicity are significantly different among sexes. Whereas smoking did not reveal significant sex-dependent differences in ruptured IAs, analysis of BP indicated that women benefit significantly more from treating hypertension with regard to IA rupture risk. Importantly, our results suggest that IAs in females have a higher endothelial coverage of the aneurysm wall and exhibit a significantly less severe histological aneurysm wall type. We propose that our observations should be taken into account for the clinical management of IAs and eventually find their way into IA management guidelines: accounting for IA and patient characteristics (which obviously influence treatment indication), aggressive treatment (neurosurgical or endovascular) in combination with reduction of risk factors might be justified in men while conservative treatment with control of risk factors (treatment of hypertension, stop smoking) might suffice in women.

Most reports on sex-dependent differences in IAs are based on cohorts of ruptured IAs alone. Ghods and colleagues^25^ reported, in a study with both ruptured and unruptured IAs including 608 patients (682 IAs), sex prevalence rates similar (72% women, 28% men) to ours and the ones found in autopsy cases (ISUIA) ^35^. Similarly, an increased frequency of multiple IAs among women is in line with earlier publications^25,36^. Regarding location and rupture risk, we identified ACOM IAs in men to have the highest rate of rupture, as previously reported^36^. Interestingly, Ghods et al.^25^ reported an increased ruptured rate in women (28% vs 23% in men), whereas we found an increased rupture rate in men (32.78% vs 25.5% in women). A recent meta-analysis by Zuurbier et al.^28^ also reported a higher risk rupture of IAs in women. Of note, in the latter study^28^, the Swiss population already appeared to be different with a higher rupture risk in men, which is consistent with our results and warrants further research. Smoking and hypertension were evaluated as risk factors/comorbidities. Smoking is considered a risk factor for both the development and rupture of IAs^37,38^. Among the patients that qualify as ex-smokers and presented with ruptured IAs, we noted that the proportion of men is significantly higher, indicating that the protective effect of smoking cessation is more important for women but still recommendable for men. Hypertension is also a known risk factor for the development and rupture of IAs^37,38^. In our results, we observed a significantly lower proportion of ruptured aneurysms among women with treated hypertension and normotension as compared to women with untreated hypertension, which was not the case in men. These results suggest that women might benefit significantly more from treating hypertension than men in regard to IA rupture risk. For sex-dependent differences regarding smoking cessation as well as treatment of hypertension, further research is warranted to better understand the biological causes of these interesting observations.

Histological classification of aneurysm wall types was shown to be a reliable marker of IA instability and rupture risk^32^. Accordingly, the higher proportions of ruptured IAs in men was reflected by an increase in wall type severity. Hormonal regulation and location-dependent hemodynamic differences might underly such sex-dependent differences, but the underlying cellular and molecular mechanisms remain obscure and require future investigations.

## Limitations

This study has a number of limitations. All registry research introduces selection bias and other inherent limitations, including potential issues with data completeness, recruitment- and enrollment strategies, and confounding variables, both known and unknown^29^. Specifically, the @neurIST database has unique limitations because of the nature of IA diseases, including underdiagnosis, heterogeneity of presentation and natural history. Indeed, a certain number of patients was excluded from further analysis due to incomplete data. Furthermore, studies reporting on interrater agreement regarding IAs diagnosis report relatively low agreement rates for IA neck diameter, characterization of IA morphology and -multiplicity^39,40^. Data derived from the registry are also likely to be enriched with patients with symptomatic IAs who seek care. Finally, potential for bias in patient selection due to lack of imaging cannot be excluded.

## Conclusion

The prevalence of IAs among women enrolled in the @neurIST cohort is significantly higher as compared to men. Women showed more often multiple aneurysms whereas men were more often diagnosed with ruptured aneurysms, corresponding to a more severe histological wall type. Sex-specific differences in aneurysm location were identified and morphologic analysis showed that the mean aneurysm neck diameter was significantly higher in ruptured IAs in men as compared to women, whereas the maximum aneurysm diameter was significantly higher in ruptured IAs as compared to non-ruptured in both sexes. Moreover, susceptibility to risk factors such as smoking and hypertension also seemed to be sex-dependent. In conclusion, these data suggest that IA morphology and wall type histology might be underlying factors for the observed sex-dependent differences in IA disease. Further research is needed to unravel the cellular and molecular mechanisms underlying these differences.

## Data Availability

Data availability
Raw data were generated at the Geneva University Hospital. Derived data supporting the findings of this study are available from the corresponding author on request.

## Acknowledgments

Data were collected starting in 2006 in the context of the EU project @neurIST and SystemsX.ch initiative AneuX evaluated by the Swiss National Science Foundation. The data is hosted by the Swiss Bioinformatics Institute in the context of the Aneurysm Data Bank. The authors would like to thank Vitor Mendes Pereira, Daniel Rüfenacht and Norman Juchler for helping with data collection, harmonization, and processing.

## Author contributions

T.W. and P.B. conceived and designed the study. P.B., T.W., M.N., P.D., M.B., A.K., P.P.M., O.G., M.J., E.A.W., H. K., K. B., R.G. and K.S. collected data. S.M. performed the histological analysis. T.W., M.N., P.C., S.M. and J.B. performed the statistical analysis. T.W., P.C., S.M. and J. B. generated the figures. T.W. wrote the paper with the help of P.C., J.B. and P.B. All authors approved the final version of the manuscript.

## Supplementary material

Supplementary material is available online.

## Supplementary figure legends

**Supplementary Figure S1**

**A-C** Average aneurysm neck diameter and rupture status according to the Koivisto chart system. The average neck diameter of ACA, MCA, ICA and VBA aneurysms did not show significant differences. **E-G** Maximum aneurysm diameter and rupture status according to the Koivisto chart system showed significantly higher values in ruptured aneurysms as compared to unruptured aneurysms for both females and males in ACA (**E**) (+46%; P < 0.0001, +49%; P<0.0001), MCA (**F**) (+61%; P < 0.0001, +142%; P<0.0001) and for females in ICA aneurysms (**G**) (+46%; P < 0.0001, respectively). **H** Maximum aneurysm diameter and rupture status of VBA aneurysms did not show significant differences. *P < 0.05, **P < 0.01, ***P < 0.001, ****P < 0.0001 (Fisher exact test).

**Supplementary Figure S2**

**A** Proportion of aneurysms by location. The proportion of ACA aneurysms was significantly higher in male as compared to female (+83%; P<0.0001) whereas aneurysms in the ophthalmic segment of the carotid artery were significantly more frequent in female as compared to male (+146%; P<0.0001). **B** Proportion of multiple aneurysms in the ophthalmic segment of the carotid artery was significantly more frequent in female as compared to male among the patients with multiple IAs (+244%; P<0.0001). **C-F** Number, proportion, aneurysm neck diameter and maximum aneurysm diameter by location were not significant between females and males. ****P<0.0001 (Fisher exact test).

**Supplementary Figure S3**

**A-F** Patients with IAs were classified, according to smoking status, into “still smoking”, “quit smoking”, and “non-smoking”. The proportion of smokers as compared to non-smokers and divided into patients with ruptured and unruptured IAs did not show significant differences in females (**A**) nor in males (**D**). The proportion of smokers as compared to patients that quit smoking, divided into patients with ruptured and unruptured IAs did not show significant differences in females (**B**) nor in males (**E**). The proportion of patient that quit smoking as compared to non-smokers, divided into patients with ruptured and unruptured IAs showed a significant difference in females (**c**) (P < 0.05) but not in males (**F**). **G-L** Patients with IAs were classified, according to blood pressure, into “no hypertension”, “treated and controlled hypertension”, “treated and uncontrolled hypertension” and “untreated hypertension”. (**G,J**) The proportion of patients with treated and controlled hypertension as compared to patients with normotension, divided into patients with ruptured and unruptured IAs showed no significant differences in females (**G**) but significant differences in males (**J**) (P < 0.05). (**H,K**) The proportion of patients with treated and controlled hypertension as compared to patients with uncontrolled hypertension, divided into patients with ruptured and unruptured IAs showed significant differences in females (**H**) (P < 0.0001) but not in males (**K**). (**I,L**) The proportion of patients with normotension as compared to patients with untreated hypertension, divided into patients with ruptured and unruptured IAs showed significant differences in females (**I**) (P < 0.001) but not in males (**L**). *P < 0.05, **P < 0.01, ***P < 0.001 (Fisher exact test).

## SUPPLEMENTARY TABLE LEGENDS

**Supplementary Table 1 Patient and intracranial aneurysm characteristics in the full cohort**

Demographical, clinical and intracranial aneurysms characteristics are listed and evaluated in both sexes.

^X^A current smoker is a patient currently smoking more than 300 cigarettes and a former smoker is a patient who smoked more than 300 cigarettes and who stopped at least 6 months ago. ^Y^Arterial hypertension is defined as blood pressure greater than 140/90 mmHg independently of the existence or not of a treatment against hypertension. Comparisons of distributions and medians have been performed using Fisher’s exact test and non-parametric Mann-Whitney *U* test, respectively.

IA= intracranial aneurysm.

**Supplementary Table 2 Aneurysm and patient characteristics of the aneurysms included in the histological wall type analysis**

Rough appearance is defined as the presence of sub-millimetric irregularities. Positive family history for IA is defined as one or more 1st degree relative(s) with IA. A current smoker is a patient currently smoking more than 300 cigarettes and a former smoker is a patient who smoked more than 300 cigarettes and who stopped at least 6 months ago. Arterial hypertension is defined as blood pressure greater than 140/90 mmHg independently of the existence or not of a treatment against hypertension. Comparisons of distributions and medians have been performed using Fisher’s exact test and non-parametric Mann-Whitney *U* test, respectively.

IQR= interquartile range; ICA= internal carotid artery; MCA= middle cerebral artery; ACA= anterior cerebral arteries, including: anterior cerebral artery, anterior communicating artery (ACOM) and pericallosal artery (A2).

