## Supplementary figures and images for "Sex-dependent manifestations of intracranial aneurysms"

### Supplementary Figure 1

#### Supplementary Figure 1

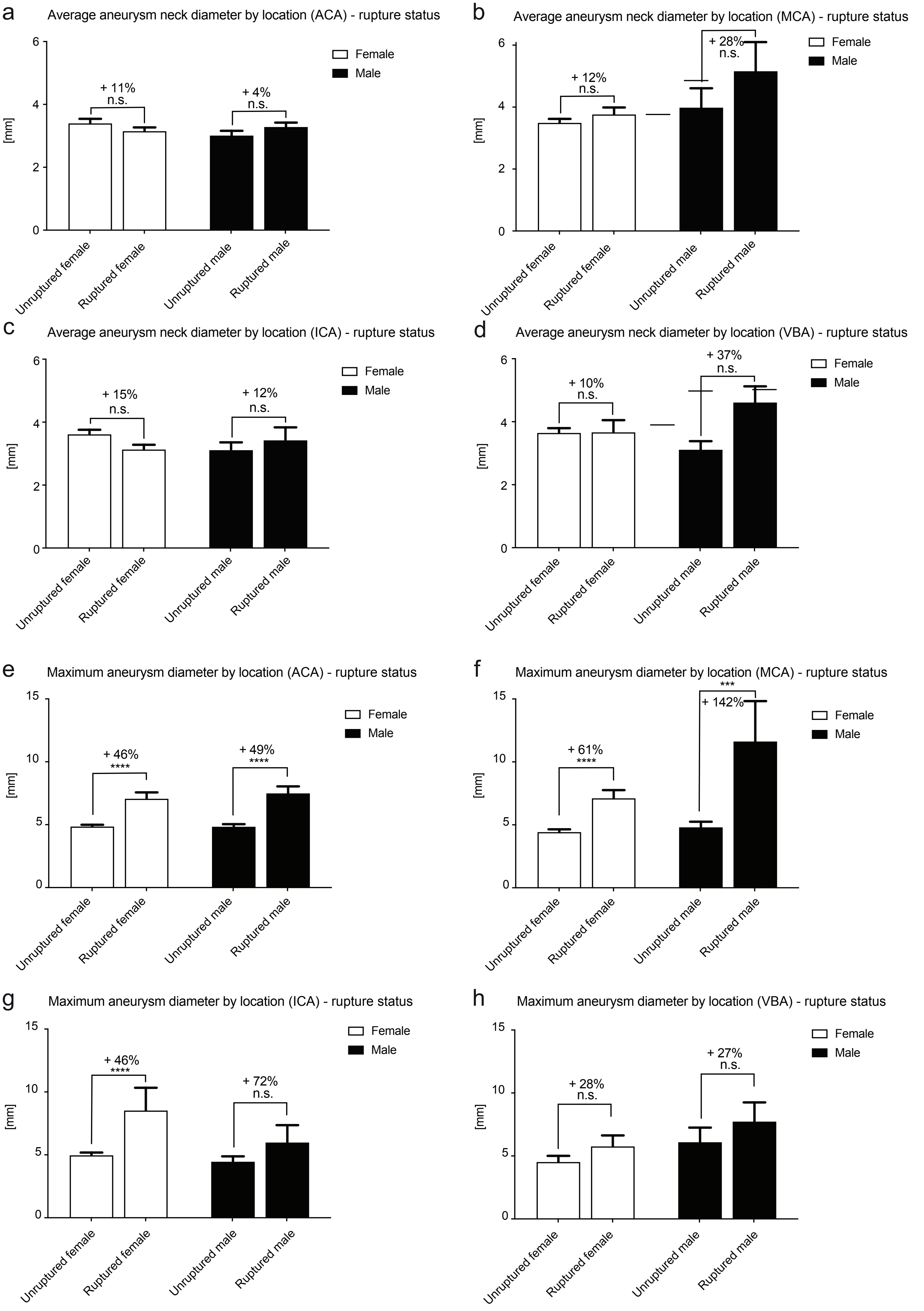

### Supplementary Figure 2

#### Supplementary Figure 2

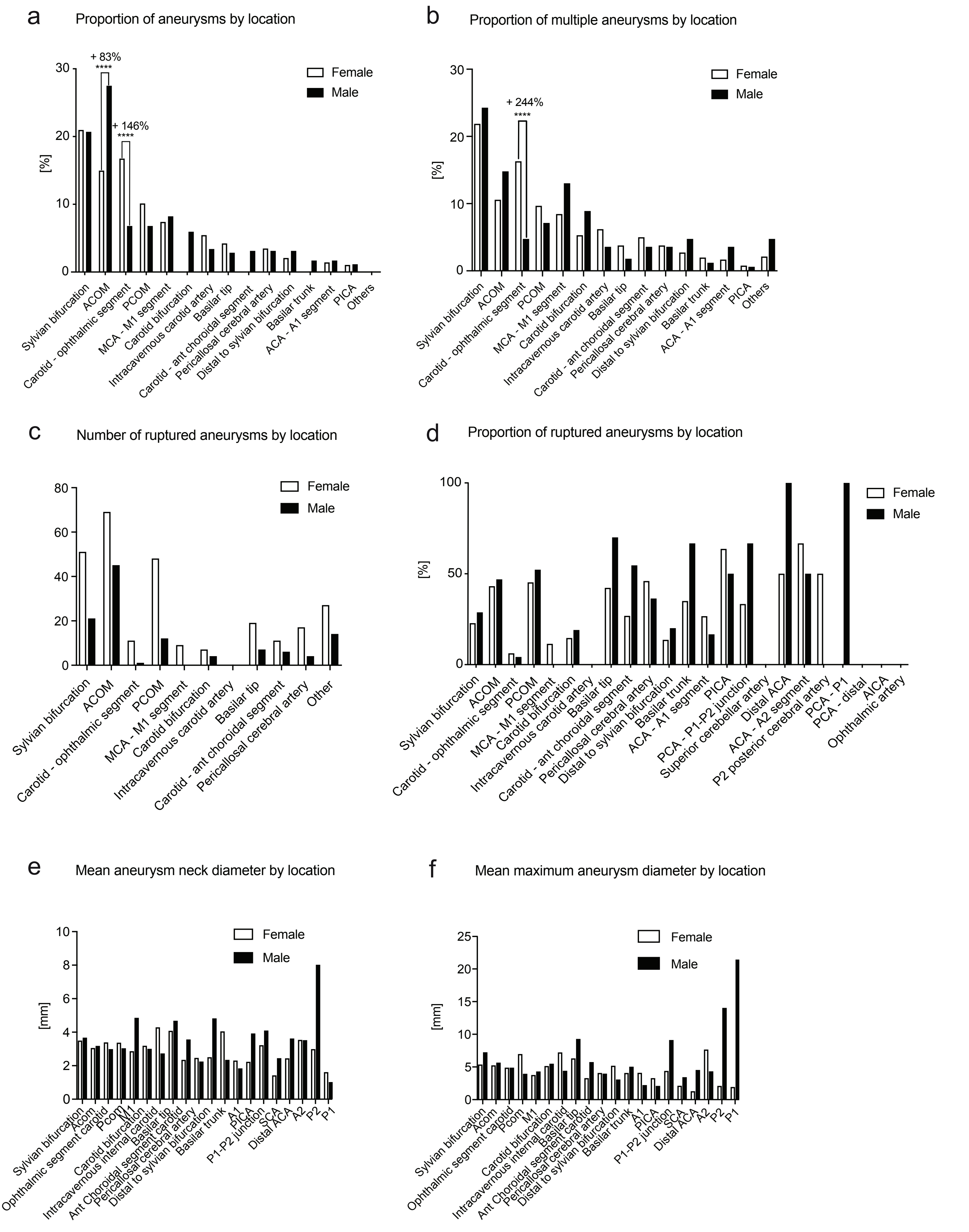

### Supplementary Figure 3

#### Supplementary Figure 3

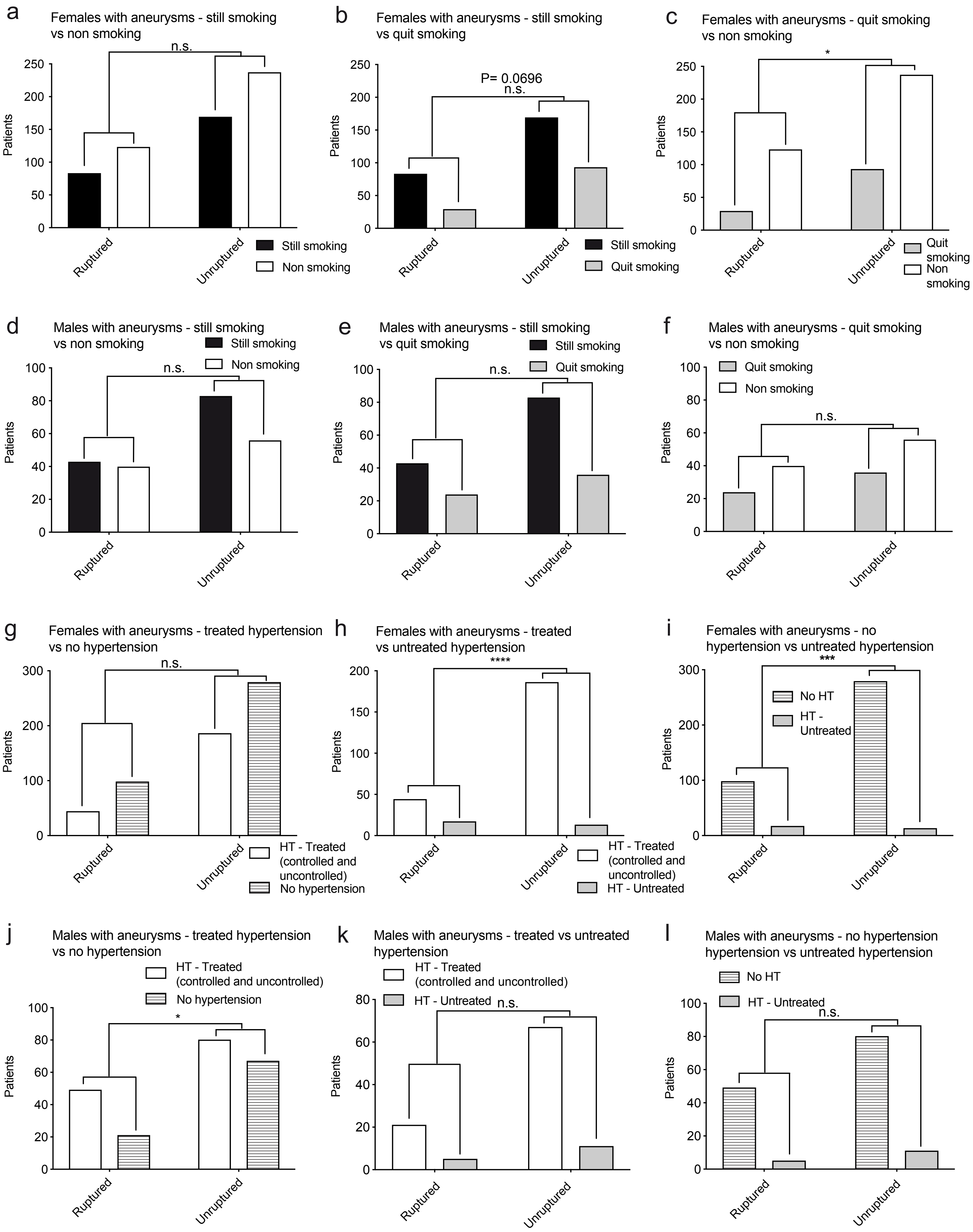
