## Supplementary Table 1 for "Sex-dependent manifestations of intracranial aneurysms"

| Characteristic | Total cohort (n=982) | Female (n=714) | Male (n=268) | P value | Statistical test |
|---|---|---|---|---|---|
| **Number of patients, n (%)** | 982 | 714 (72.7) | 268 (27.3) | **<0.001** | Chi-square |
| **Number of IAs, n (%)** | | | | | |
| Unruptured IAs | 1056 (72.7) | 812/1090 (74.5) | 244/363 (67.2) | **<0.001** | Chi-square |
| Ruptured IAs | 397 (27.3) | 278/1090 (25.5) | 119/363 (32.8) | **0.008** | Chi-square |
| **Age at diagnosis, y (median, SD)** | 55.8 (13.96) | 56.1 (13.39) | 55.6 (14.06) | >0.05 | Mann-Whitney |
| **Medical history, n (%)** | | | | | |
| Subarachnoid hemorrhage (IA rupture) | 397 (27.3) | 278/1090 (25.5) | 119/363 (32.8) | **0.008** | Chi-square |
| Multiple IAs | 363 (37.0) | 289/714 (40.5) | 74/268 (27.6) | **<0.001** | Chi-square |
| Tobacco (current[x]) | 329 (33.5) | 220/714 (30.8) | 109/268 (40.6) | >0.05 | Chi-square |
| **IA dimensions, mm (median, SD)** | | | | | |
| Average IA neck diameter – Unruptured | | 3.409 (2.155) | 3.467 (3.891) | >0.05 | Mann-Whitney |
| Average IA neck diameter – Ruptured | | 3.488 (1.76) | 3.91 (2.853) | >0.05 | Mann-Whitney |
| Maximum IA diameter – Unruptured | | 4.956 (3.887) | 4.905 (3.689) | >0.05 | Mann-Whitney |
| Maximum IA diameter – Ruptured | | 7.571 (9.491) | 8.131 (8.108) | >0.05 | Mann-Whitney |
