## Supplementary Table 2 for "Sex-dependent manifestations of intracranial aneurysms"

| Characteristic | Female (n=36) | Male (n=12) | *P* value | Statistical test |
|---|---|---|---|---|
| **Number of patients, n (%)** | 36 (75) | 12 (25) | | |
| **Age, mean (SD), y** | 53 (11) | 56 (11) | 0.38 | Mann-Whitney |
| **Medical history, n (%)** | | | | |
| Previous aSAH | 4 (11) | 1 (9) | 1 | Fisher's exact |
| Multiple IAs | 21 (62) | 5 (45) | 0.48 | Fisher's exact |
| Tobacco (former and current) | 20 (57) | 10 (91) | 0.07 | Fisher's exact |
| Hypertension | 16 (46) | 4 (36) | 0.73 | Fisher's exact |
| Family history of IAs | 8 (21) | 0 (0) | 0.17 | Fisher's exact |
| **IA characteristics** | | | | |
| Ruptured IAs, n (%) | 14 (39) | 3 (33) | 1 | Fisher's exact |
| Location, n (%) | | | | |
| *MCA* | 27 (75) | 8 (67) | 0.52 | Fisher's exact |
| *ACA* | 6 (16.5) | 4 (33) | | |
| *ICA* | 2 (5.5) | | | |
| *Unknown* | 1 (3) | | | |
| Maximum IA diameter, mm, median (IQR) | 6.8 (5.4-8.0) | 5.9 (4.0-11.5) | 0.93 | Mann-Whitney |
| Average IA neck diameter, mm, median (IQR) | 3.6 (3.0-4.4) | 3.5 (2.0-6.0) | 0.93 | Mann-Whitney |
| Rough appearance, n (%) | 13 (43) | 5 (42) | 1 | Fisher's exact |
| Presence of blebs/lobules, n (%) | 20 (59) | 7 (64) | 1 | Fisher's exact |
